# Seroprevalence of novel coronavirus disease (COVID-19) in Kobe, Japan

**DOI:** 10.1101/2020.04.26.20079822

**Authors:** Asako Doi, Kentaro Iwata, Hirokazu Kuroda, Toshikazu Hasuike, Seiko Nasu, Aya Kanda, Tomomi Nagao, Hiroaki Nishioka, Keisuke Tomii, Takeshi Morimoto, Yasuki Kihara

**Affiliations:** Department of Infection Control, Kobe City Medical Center General Hospital, Chuoku 6500047, Kobe, Hyogo, Japan; Department of Infectious Diseases, Kobe City Medical Center General Hospital, Chuoku 6500047, Kobe, Hyogo, Japan; Division of Infectious Diseases Therapeutics, Kobe University Graduate School of Medicine, Chuoku 6500017, Kobe, Hyogo, Japan; Department of Laboratory Medicine, Kobe City Medical Center General Hospital, Chuoku 6500047, Kobe, Hyogo, Japan; Department of Respiratory Medicine, Kobe City Medical Center General Hospital, Chuoku 6500047, Kobe, Hyogo, Japan; Department of Clinical Epidemiology, Hyogo Medical College, Mukogawa 6638501, Nishinomiya, Hyogo, Japan; Kobe City Medical Center General Hospital, Chuoku 6500047, Kobe, Hyogo, Japan

## Abstract

**Background:** Coronavirus disease 2019 (COVID-19) pandemic caused by SARS-CoV-2 has been affecting many people on earth and our society. Japan is known to have relatively less number of infections as well as deaths among developed nations. However, accurate prevalence of COVID-19 in Japan remains unknown. Therefore, we conducted a cross-sectional study to estimate seroprevalence of SARS-CoV-2 infection.

**Methods:** We conducted a cross-sectional serologic testing for SARS-CoV-2 antibody using 1,000 samples from patients at outpatient settings who visited the clinic from March 31 to April 7, 2020, stratified by the decade of age and sex.

**Results:** There were 33 positive IgG among 1,000 serum samples (3.3%, 95%CI: 2.3-4.6%). By applying this figure to the census of Kobe City (population: 1,518,870), it is estimated that the number of people with positive IgG be 50,123 (95%CI: 34,934-69,868). Age and sex adjusted prevalence of positivity was calculated 2.7% (95%CI: 1.8-3.9%), and the estimated number of people with positive IgG was 40,999 (95%CI: 27,333-59,221). These numbers were 396 to 858 fold more than confirmed cases with PCR testing in Kobe City.

**Conclusions:** Our cross-sectional serological study suggests that the number of people with seropositive for SARS-CoV-2 infection in Kobe, Japan is far more than the confirmed cases by PCR testing.

## Introduction

Coronavirus disease 2019 (COVID-19) pandemic caused by SARS-CoV-2 has been affecting many people on earth and our society. As of the middle of April,2020, Japan is known to have relatively small number of infections as well as deaths among developed nations. As of April 25, 2020, only 12,240 infections were identified with 317 death.^1^ However, with restricted use of polymerase chain reaction (PCR) testing to diagnose COVID-19 in Japan, some might be skeptical about the figure, with potential many undiagnosed people without PCR testing.^2^ Accurate prevalence of COVID-19 in Japan remains unknown and many are concerned about this uncertainty. To estimate actual number of infected persons in community, we conducted a cross-sectional study to estimate seroprevalence of SARS-CoV-2 infection.

## Methods

We conducted a cross-sectional serologic testing for SARS-CoV-2 antibody (immunoglobuin G, or IgG) at Kobe City Medical Center General Hospital, a tertiary care medical center in Kobe, Japan. Tests were done for randomly selected preserved serum from patients who visited outpatient clinics of the hospital and received blood testing for any reason. Patients who visited the emergency department or the designated fever consultation service were excluded to avoid the overestimation of SARS-Cov-2 infection. The serums of patients who visited the center from March 31 to April 7 were used for the analysis. Serums were preserved at −20 and were defrosted upon testing. An immunochromatographic test to detect IgG against SARS-CoV-2 was used for the analysis (RCNC002, KURABO Industries Ltd. https://www.kurabo.co.jp/bio/English/index.html). Ten microliter of serum was infused into the test kit, and the interpretation of the test results were made 15 minutes after the buffer was placed. Tests were conducted for 1,000 samples, stratified by the decade of age and sex, with 50 samples taken from each stratum. If there were less than 50 samples from any stratum, additional samples were randomly obtained from other strata until all 1,000 samples were completed. This study was conducted anonymously with exception of age groups and completely impossible. The estimate of this study was prevalence of seropositivity of IgG and its 95% confidence interval (CI). Both unadjusted and age and sex adjusted estimations of number of seropositive persons in Kobe city were also calculated. For unadjusted estimation, we used Kobe City website for population of Kobe City (of April 1, 2020 https://www.city.kobe.lg.jp/a89138/shise/toke/toukei/jinkou/index.html). For age and sex adjusted estimation, we used data of national census conducted in 2015 (https://www.city.kobe.lg.jp/a89138/shise/toke/toukei/kokutyou/tyoubetsujinkou.html). The number of confirmed cases as of April 7, 2020 was 69 and used based on the data presented by Kobe City (https://www.city.kobe.lg.jp/a57337/kenko/health/coronagaiyo1_100.html).

The ethics committee at Kobe City Medical Center General Hospital approved the current study.

## Results

There were 33 positive IgG among 1,000 serum samples (3.3%, 95%CI: 2.3%-4.6%). By applying the estimate to census of Kobe City (population: 1,518,870), it is estimated that the number of people with positive IgG be 50,123 (95%CI: 34,934-69,868). Since total number of infected patients reported at Kobe City as of April 7 were only 69, these estimates were 506 to 1,013 fold more than confirmed cases then.

Sample characteristics are shown on Table 1 stratified by the decade of age and sex of the patients. Table 2 shows age and sex distribution of Kobe City at the national census held in 2015. The age and sex adjusted percentage of positivity was 2.7% (95%CI: 1.8-3.9%). Using calculated population of Kobe City in national census of 2015 (1,518,478), estimated number of people with positive IgG was 40,999 (95%CI: 27,333-59,221). These numbers were 396 to 858 fold more than confirmed cases in Kobe City.

**Table 1.** Sample characteristics

| Ages | Male | Test positive | Female | Test positive |
| --- | --- | --- | --- | --- |
| Under 10-year-old | 7 | 0 | 1 | 0 |
| 10-19 | 17 | 0 | 10 | 0 |
| 20-29 | 17 | 0 | 19 | 0 |
| 30-39 | 41 | 1 | 49 | 2 |
| 40-49 | 64 | 2 | 91 | 3 |
| 50-59 | 84 | 4 | 80 | 2 |
| 60-69 | 87 | 2 | 84 | 3 |
| 70-79 | 85 | 6 | 81 | 3 |
| 80-89 | 81 | 1 | 83 | 4 |
| Over 90 | 6 | 0 | 13 | 0 |
| Total | 489 | 16 | 511 | 17 |

**Table 2.** Population of Kobe City based on 2015 census. Total number of the populations are aggregates of all age groups, which are different from what the census figure showed.

| Ages | Male (%) | Female (%) |
| --- | --- | --- |
| Under 10-year-old | 61,242 (8.6) | 58,671 (7.3) |
| 10-19 | 70,275 (9.8) | 67,661 (8.4) |
| 20-29 | 73,973 (10.3) | 78,787 (9.8) |
| 30-39 | 87,806 (12.3) | 95,510 (11.9) |
| 40-49 | 109,303 (15.3) | 116,372 (14.5) |
| 50-59 | 89,500 (12.5) | 98,220 (12.2) |
| 60-69 | 105,160 (14.7) | 114,649 (14.3) |
| 70-79 | 77,705 (10.9) | 96,228 (12.0) |
| 80-89 | 36,428 (5.1) | 61,344 (7.6) |
| Over 90 | 4,475 (0.6) | 15,169 (1.9) |
| Total | 715,867 | 802,611 |

## Discussion

Our cross-sectional serological survey identified far more patients than those actually diagnosed by PCR testing. The prime minister of Japan Shinzo Abe declared the state of emergency in the afternoon on April 7, 2020, and it means that samples of our analysis reflect the status before the state of emergency.^3^

Japan’s government tried to restrict the number of PCR testing to avoid a collapse of health care system in Japan, and only those who with persisting symptoms or those with risk factors were allowed to take PCR. It was considered that identifying clusters occurring in Japan with monitoring and isolating close contacts with the identified patients were prudent strategy and it would be able to comprehend the trend of COVID-19 infections in Japan.^4^ However, since difference between those who were identified and those who actually had infections was so huge according to our findings, Japan might not have comprehended the epidemic dynamics occurring in the nation.

The thought that confirmed COVID-19 cases were a tip of iceberg is not new. A similar study by Bendavid *et al*. identified that seroprevalence of SARS-CoV-2 at Santa Clara County, the United States, was 1.5%, and the estimated number of cases were 50-85 fold more than confirmed cases.^5^ A previous study tried to estimate actual cases of COVID-19 and it concluded that close to half the cases are identified in Japan.^6^ We consider this report underestimated the actual infection rate and excessively overestimated the ability of Japan’s capability of diagnosing COVID-19. The study on the outbreak of COVID-19 in a cruise ship Diamond Princess suggested that there are about 20-40% asymptomatic cases among those infected with SARS-CoV-2,^7^ and this could suggest finding of every case of COVID-19 rather unrealistic. However, even with this assumption, missing cases by several hundred fold is not acceptable, and better strategy should have been implemented to better fight against this infection.

Because of the low number of confirmed cases in Japan, there was discussion whether Japan is different from other nations in terms of transmissibility of SARS-CoV-2. Our analysis suggests that the transmissibility of the virus in Japan might not differ significantly from other countries. This means the difference in behavior of Japanese people, such as lack of hugs or kissing in public, custom of taking off shoes at the entrance, or wearing of masks for protection purpose before the outbreak, might not be the reason. Rather, Japan probably had significant transmission of SARSCoV-2 like other countries. On the other hand, is the data on mortality due to COVID-19 reliable in Japan? There is no thorough investigation on the accuracy of mortality data regarding COVID-19 in Japan, but concordance of death certificates and autopsy diagnoses for pneumonia in Japan was only 9%, and the possibility of underestimates of mortality also needs to be considered.^8^ If we consider the mortality data in Japan accurate, it raises another question why there is fewer deaths due to COVID-19 in Japan. Vaccination of BCG (Bacille de Calmette et Guerin) are hypothesized to affect the clinical impact of COVID-19,^9^ but further studies will be needed to confirm its effects.

Our study has several inherent limitations. First, the sensitivity and specificity of serology assay to estimate the prevalence of SARS-CoV-2 infection is not fully studied. There might be underdiagnosis of the viral infection, particularly at the early stage of infection, or those with immunosuppressed. The possibility of overdiagnosis by cross-reaction with other viruses, such as conventional coronavirus as a cause of common cold, although it had never been documented as of this writing, should be considered. Second, the population of our cohort might not be the same as one in Kobe City. The samples were obtained from the patients from the clinic. Although we adjusted the age and sex distribution to fit the general population, the selection bias should be unavoidable. Further serological studies targeting randomly selected people in Kobe City could clarify this potential limitation. However, even we put these potential limitations into consideration, the difference between what had been reported as the confirmed cases and estimated cases in our study is so huge, and our interpretation of the findings sounds rather robust and valid.

In conclusion, our cross-sectional serological study suggests that the actual number of people with SARS-CoV-2 infection in Kobe, Japan is far more than the confirmed cases by PCR testing. Further studies is needed to elucidate the seroprevalence in other regions in Japan and the dabate regarding lower mortality in Japan.

## Data Availability

Data is available upon request.

## The Contributors

AD, KI, and TM developed idea and design of the study. All contributed equally to review and finalize the study design. AD, KK, TH, SN, AK, and TN collected data and conducted serology tests from preserved samples. AD, KK, TH, KI, and TM participated in data interpretation and statistical analysis. AD and KI wrote the draft of manuscript, and KK, TH, SN, AK, TN, HN, KT, and TM critically revised the manuscript.

## Author approval

All authors have seen and approved the manuscript.

## Data availability statement

Data is available upon request.

## Acknowledgements

We thank Ms. Toko Nakata and Mako Miyawaki for aiding the purchase of test kits.

